## Supplementary Files (1-4) for "Use of Generative AI for Health Among Urban Youth in Pakistan: A Mixed-Methods Study"

**Supplement 1**

The screencap (Figure S1) attached below demonstrates (1) the increase in searches of "ChatGPT", a GAI tool, (2) and the narrowing gap of this keyword compared with "Google". This screencap was taken in June 2025, and shows 12 months i.e., June 2024 to June 2025.

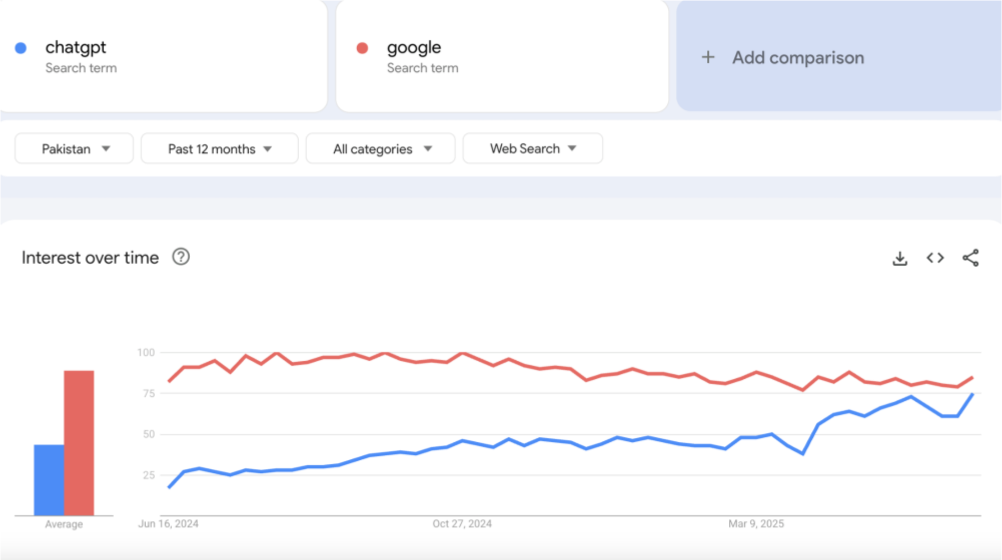


**Figure S1.1: Search Terms Google vs. ChatGPT in Pakistan via Google Trends**

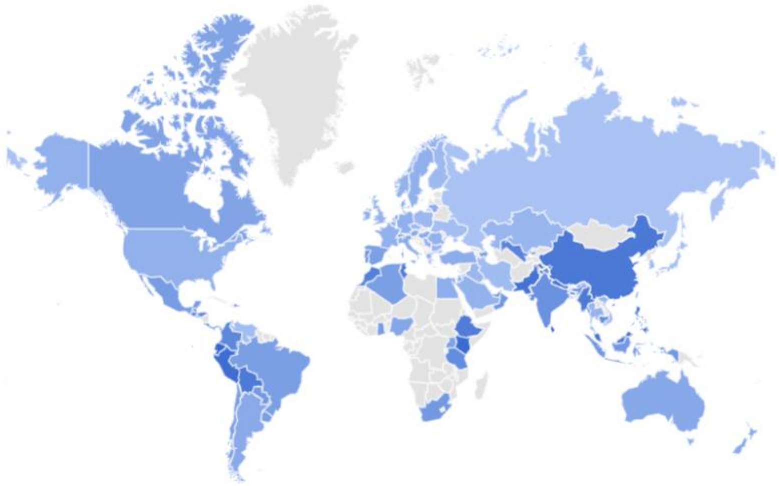

**Figure S1.2: Worldwide Search Results for "ChatGPT" by country via Google Trends**

Listed below are the questions asked in the quantitative survey tool of this research study, followed by a list of variables categorized by each section.

### CONSENT

**Consent:** By clicking 'Yes,' I confirm that I am 18–30 years old and voluntarily agree to participate in this anonymous study on AI and health.

### SECTION I: ABOUT YOU

**Q1:** What is your age?

**Q2:** What is your gender?

**Q3:** Which of the following best describes your sexual orientation?

**Q4:** Which of the following boards did you study from?

**Q5:** How many people in your life would you consider close friends or companions you trust?

**Q6:** Do you feel comfortable discussing sensitive or personal health concerns with your family?

**Q7:** Do you have any existing or past medical or mental health conditions (diagnosed or undiagnosed)?

**Q8:** Have you ever used any of the following non-generative AI or tech-enabled health tools? (Select all that apply)

**Q9:** How often do you use AI tools like ChatGPT, DeepSeek, Gemini, etc?

### SECTION II: USE OF GENERATIVE AI FOR HEALTH

**Q1:** Have you ever used a generative AI platform to ask about a health-related concern?

**Q2:** Which AI platform(s) have you used for health-related questions? (Select all that apply.)

**Q3:** When you've used AI for health-related concerns, who was it for?

**Q4:** What types of health issues have you asked about using AI? (Select all that apply.)

**Q5:** What first led you to try an AI platform for a health-related concern? (Select all that apply)

**Q6:** How helpful did you find the AI's response to your health concern?

**Q7:** How frequently do you use AI for health-related concerns?

**Q8:** What do you usually do after receiving health-related advice from an AI platform?

**Q9:** Compared to visiting a doctor, how comfortable do you feel asking sensitive health questions to an AI platform?

### SECTION III: TRUST IN AI

**Q1:** How much do you/would you trust the information provided by AI platforms like ChatGPT for health-related questions?

**Q2:** How confident are you about using AI platforms to find and understand health-related information?

**Q3:** Are you aware of any potential risks of using AI platforms, either in general or for health-related concerns?

**Q4:** Which of the following are you concerned about regarding usage of AI for health? (Select all that apply)

### SECTION IV: ACCESS TO HEALTHCARE

**Q1:** If you needed to see a doctor today, how easy would it be for you to arrange that on your own?

**Q2:** Overall, how satisfied are you with the healthcare services available to you?

**Q3:** How often does getting professional healthcare for a health concern end up being delayed or avoided in your case?

**Q3b:** What are the most common reasons why you have delayed or avoided seeking healthcare? (Select the top 3 options)

### LUCKY DRAW ENTRY

**Optional:** Would you like to enter a lucky draw for a small token of appreciation.

**Section 1 variables:** `age`, `gender`, `sexual_orientation`, `education_board`, `num_close_friends`, `comfortable_sharing_health_with_family`, `existing_health_conditions`, `non_generative_health_tool_use`, `ai_tool_usage_frequency`

**Section 2 variables:** `used_ai_for_health`, `ai_platforms_used_for_health`, `who_used_ai_for`, `health_issues_asked_about`, `reason_for_using_ai_for_health`, `ai_response_helpfulness`, `frequency_of_ai_use_for_health`, `action_after_ai_advice`, `comfort_comparing_ai_vs_doctor`

**Section 3 variables:** `trust_in_ai_for_health`, `confidence_in_using_ai_for_health`, `aware_of_ai_health_risks`, `ai_health_concerns`

**Section 4 variables:** `ease_of_arranging_doctor_visit`, `satisfaction_with_healthcare_services`, `frequency_of_delaying_healthcare`, `reasons_for_delaying_healthcare`

Supplement 3

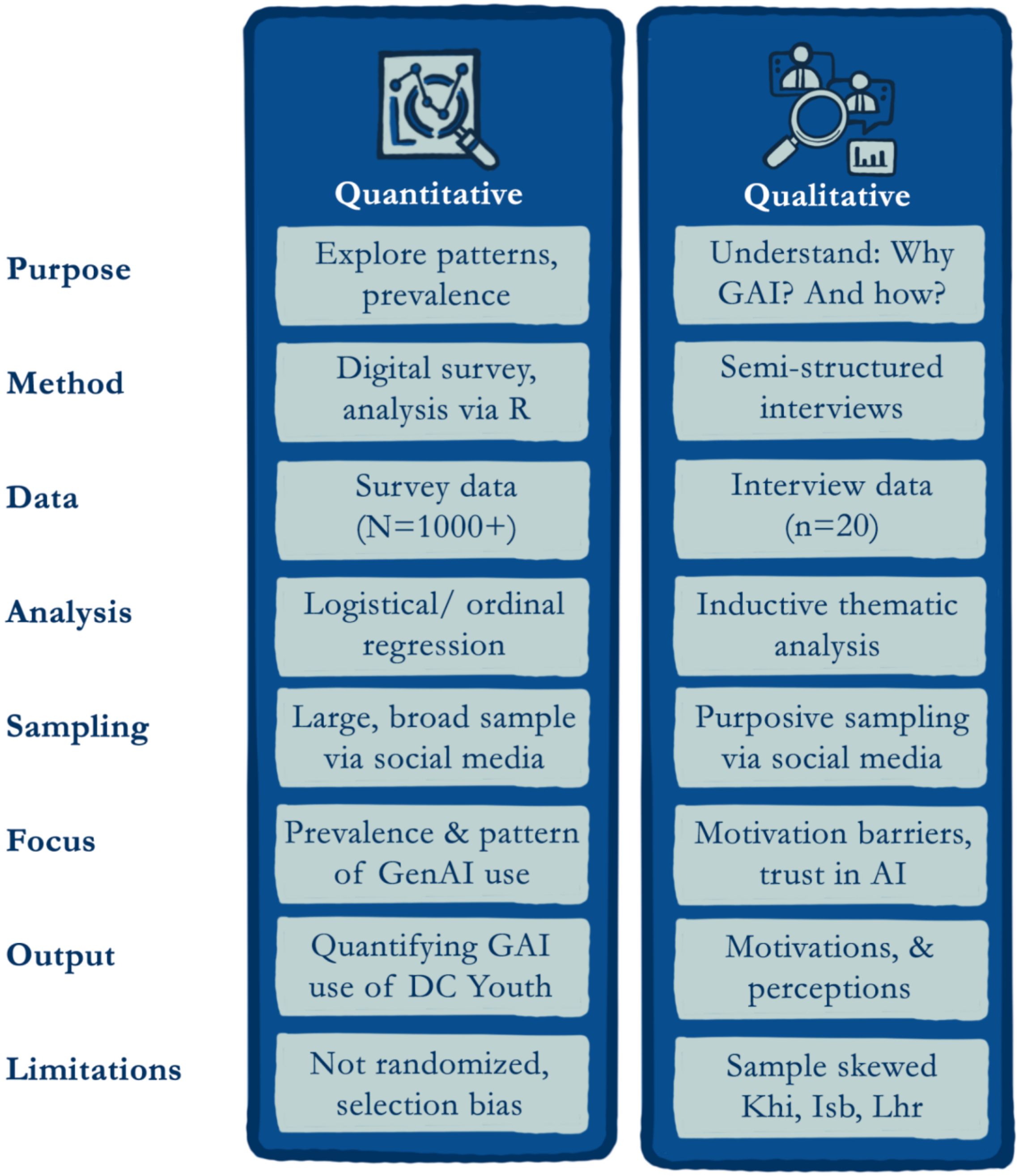


**Figure S2: Good Reporting of A Mixed-Methods Study Checklist (O'Cathain et al., 2008)**

**Supplement 4**

Notes: This checklist follows the 32 COREQ items. Responses reflect the qualitative component (n=20 interviews) using reflexive thematic analysis.

| Item # | COREQ item | Response for this study |
|---|---|---|
| 1 | Interviewer/facilitator | All five authors (AM, AA, IK, MH, SB) conducted interviews. |
| 2 | Credentials | Undergraduate/early-career researchers (ages 20–30) at Habib University; trained in qualitative methods and YPAR. AM (MSc, Incoming DPhil), AA, IK, MH, SB (BSc Social Development Policy) |
| 3 | Occupation | Students/research fellows at Habib University during the study. |
| 4 | Gender of interviewer | Mixed team (women and men). |
| 5 | Experience/training | Team training in semi-structured interviewing, reflexive TA; piloting and cognitive testing conducted. |
| 6 | Relationship established | Minimal/No prior relationships with participants. |
| 7 | Participant knowledge of interviewer | Participants informed the study was youth-led, academic, non-profit; aims, roles, and confidentiality explained in consent. |
| 8 | Interviewer characteristics/reflexivity | Pakistani youth researchers; digitally connected; kept reflexive memos to bracket assumptions. |

| | | |
|---|---|---|
| 9 | Methodological orientation | Reflexive Thematic Analysis (Braun & Clarke) with an inductive approach; socio-ecological model guided inquiry. |
| 10 | Sampling | Purposive/self-selection of urban youth (18–30) who reported routine GAI use for health. |
| 11 | Method of approach | Social media recruitment (stories/posts); scheduling via messages/email; virtual interviews (Trello). |
| 12 | Sample size | n = 20 interviews. |
| 13 | Non-participation | Not systematically recorded; some initial contacts did not schedule or later withdrew due to availability. No formal refusals documented. |
| 14 | Setting of data collection | Virtual (Zoom/Google Meet) to maximize geographic access and comfort. |
| 15 | Presence of non-participants | None. |
| 16 | Sample description | Urban young adults (18–30), gender-mixed; digitally connected; routine GAI users; pseudonyms assigned. |
| 17 | Interview guide | Semi-structured guide developed from formative work; piloted and refined. (Guide available on request; domains reported in Methods 2.2.) |
| 18 | Repeat interviews | None. |
| 19 | Audio/visual recording | Yes: audio-recorded with consent. |

| | | |
|---|---|---|
| 20 | Field notes | Yes: taken during and after interviews. |
| 21 | Interview duration | ~30–60 minutes. |
| 22 | Data saturation | Not used; adequacy judged via information power: later interviews added nuance, not new themes. |
| 23 | Transcript return | No transcript/member checking (to preserve anonymity and minimize burden). |
| 24 | Number of data coders | Team-based analysis; AA. IK MH SB. AM all coded. |
| 25 | Coding tree description | Inductive codebook iteratively developed and refined via team discussions and memoing; available on request. |
| 26 | Theme derivation | Inductive; themes developed through reflexive engagement with data. |
| 27 | Software | Mixed: Turboscribe for transcription; shared docs/spreadsheets and memoing for analysis (no dedicated CAQDAS); R used for quant only. |
| 28 | Participant checking of findings | No (see item 23 rationale). |
| 29 | Quotations presented | Yes—illustrative quotes with pseudonyms and basic descriptors included. |
| 30 | Data–finding consistency | Clear alignment between quotes, codes, and reported themes. |
| 31 | Clarity of major themes | Three major themes reported (Access/affordability; Emotional safety/support; Empowerment/agency). |

| | | |
|---|---|---|
| 32 | Clarity of minor themes | Minor/divergent signals noted (e.g., climate anxiety; mixed signals on prior tool use). |
